# Health perception among the elderly in India: A gender perspective

**DOI:** 10.1101/2021.12.18.21268022

**Authors:** Saddaf Naaz Akhtar, Nandita Saikia

## Abstract

There is an urgent need to understand and study the gender-based comprehensive own-perception approaches about health status. Our primary interest is to elucidate and capture whether and what determinants of gender disparity exist in own-perception about current and change in health status in Indian settings among the elderly. Therefore, we intended to examine the gender disparity in own-perception and their differences among the elderly in India. We used cross-sectional data from the 75^th^ National Sample Survey Organizations (NSSO), collected from July 2017 to June 2018. The analytical sample constitutes 42759 cases of the elderly (eliminating two points of transgender). Thus, 21902 older men and 20857 older women have been considered. Two different measurements of own-perception about health status among the elderly have been used. We have calculated absolute gaps in the prevalence of current self-perception and change in health status by background characteristics. We carried out an ordered logistic regression model (or proportional odds model) to determine the predictors of health perception among the elderly. There is a clear gap between men and women in terms of rating poor perception about health; men generally have reported higher perception about their current health status when it comes to rating them excellent in terms of socio-economic outcomes like income, place of residence, and household structure. Despite numerous limitations, this study addressed the significant public health concern, which is crucial to address the challenge of the elderly health and their perception of well-being.

## Introduction

Aging is an unavoidable process in physiological terms. According to the World Health Organization (2020), the populations around the world are aging faster than in the past, and its demographic transition would have a significant impact on almost all aspects of society (WHO, 2020). Every country throughout the world is experiencing growth in both the proportion and size of the elderly in the population (WHO, 2021). The primary care of the elderly is mainly influenced by health services, health conditions, and socio-economic factors (Nair et al., 2021). Though, gender accentuates a pivotal role in aging with significant gaps and variations in the health conditions and the care received. Hence, the gender gap in the aging process is putting forward many health challenges and opportunities that need to be addressed. Indeed, aging healthy and successfully is an overall goal for the elderly people, policymakers, and other health professionals. Despite that, whether “healthy aging” and “successful aging” truly signify by gender among the elderly themselves is still found to be unclear. Therefore, this question can be explained by assessing self-rated health, which is the easiest way to reveal the current and change in the health status.

Own-perception about health status (OPHS) is one of the most frequent indicators used in social, clinical, and epidemiological research. It is a comprehensive measure of an individual’s health status that can even reflect their condition without any clinical diagnosis (Kananen et al., 2021). Despite its non-explicit nature, it seems to be a robust predictor of future functional and physical health status, morbidity, and mortality that may differ by gender, age, place, health status, social class, culture, and countries (Idler & Benyamini, 1997; Jylhä, 2009). Various disease risks screening (My, 2006) and clinical trials (Fayers & Sprangers, 2002) have been performed using self-rated health as a tool in developed countries. Perception about health status is an individual’s subjective concept which lies between the social & biological world with psychological experiences. Generally, the empirical research on perception about own health status arrived from the epidemiological tradition that particularly emphasized statistical associations of correlates instead of the process from which these correlations become known (Jylhä, 2009).

A large body of literature concerning OPHS has been suggested to explain its determinants and outcomes, which may reflect ill-health behavior (Garrity et al., 1978; Lee et al., 2012; Leinonen et al., 2001; Singh et al., 2013; Zimmer et al., 2000). Apart from this, OPHS also reflects psychosocial or lifestyle conditions (Harrington et al., 2010). A previous study conducted in Thailand has revealed that self-assessed health is a robust indicator of functional status, chronic diseases, and psychosocial symptoms among the elderly (Haseen et al., 2010). Another study has suggested that the elderly involved in activities of daily living limitations, worse chronic & mental health conditions, poorer self-reported memory have lower self-rated health in the United States and China (Xu et al., 2019). Despite that, previous research conducted in Taiwan has found psychological and physical propensities associations with poor SRH among the elderly (Lee et al., 2012). A longitudinal study revealed that the elderly’s physical and functional activities had been the strong predictors in self-assessments of health in Finland (Leinonen et al., 2001).

While the general public health and well-being among the Indian population have been challenging, the health disparities between older men and women have not reduced significantly. However, few studies have been conducted in India on own-perception about health status from a gender perspective (Bora & Saikia, 2015; Kumar & Pradhan, 2019; Singh et al., 2013). Earlier studies showed that gender impacts unhealthy and healthy lifestyles perceptions and gender gaps exist during health-related decision-making. Still, own-perception about health status by gender is unknown and difficult to understand because of the paucity of empirical research from both the aspects of theoretical and conceptual vagueness.

To our best knowledge, no research has been yet performed on current and changes together in health perception by gender in India among the elderly. Moreover, we already know that population aging is an emerging issue for India with significant socio-demographic, economic, and public health implications. It is an inescapable process that faces various health challenges. Older men and women present an attractive comparative setting to the medical practitioners, policymakers, and researchers to recognize the factors influencing the elderly’s health perception to identify illness behavior (Garrity et al., 1978). At the same time, the comprehensive understanding of the elderly’s health perception is poorly understood concerning gender.

There is an urgent need to understand and study the gender-based comprehensive self-perception approaches about health status. In the present study, our main interest is to elucidate and capture whether and how gender disparity exists in own-perception about current and change in health status in Indian settings among the elderly. The gender gap in health perception among the elderly is still largely unclear; however, no previous studies have attributed these gaps to the current and change in health perceptions among the elderly people in the Indian context. Therefore, the present study has intended to examine the gender disparity in own-perception and their change among the elderly in India.

## Methods

### Data source

The present study has used the data from the 25^th^ schedule of the 75^th^ round of the National Sample Survey Organizations (NSSO), collected from July 2017 to June 2018. The NSSO has been a public organization since 1950 under the Ministry of Statistics and Programme Implementation (MOSPI) of the Government of India. It is a nationally and state/Union Territory (UT) representative household, cross-sectional, population-based survey. This data is publicly published and can be accessible using https://www.mospi.gov.in/web/mospi/download-tables-data/-/reports/view/templateTwo/16202?q=TBDCAT

#### Analytical sample

The analytical sample constitutes 42759 cases of the elderly (eliminating two cases of transgender). Thus, 21902 older men and 20857 older women have been considered.

### Outcome variables

The study has used two different measurements of own-perception about health status among the elderly. Thus, two outcome variables have been used.

○ The first outcome variable is the trichotomous variable of own-perception about current health status. During the survey, the respondent asked the question to rate the individual’s perception about the current status of health using the scales. The scales were categorized into three. *Poor as ‘1’, good/fair is taken as ‘2’ and ‘Excellent/very good is taken as ‘3’*.
○ The second outcome variable is also a trichotomous variable of own perception about change in the state of health. The questionnaire has been asked the respondent to rate the individual’s own perception change in the state of health using the scales. The scales were categorized into five, but we have converted them into three in the present study (See variable description in the Supplemental files). *Somewhat worse/worse as ‘1’, nearly the same is taken as ‘2’, and* c*ompared to the previous year: much better/ somewhat better is taken as ‘3’*.

### Independent variables

Literature suggests that there are several predictors of own-perception such as social, demographic, physical, cognitive, and mental factors that have been used in Indian settings and other countries as well (Bora & Saikia, 2015; Galenkamp et al., 2013; Kumar & Pradhan, 2019; Lee et al., 2012; Leinonen et al., 2001; Singh et al., 2013; Wagner & Short, 2014). Therefore, the independent variables were applied in the present study mainly emphasized on socio-demographic & economic background characteristics and health information of the elderly. These background characteristics comprise of age groups, regions, place of residence, education level, marital status, household members, wealth quintiles, social groups (caste groups), religion, owned house, health insurance support, living arrangements, economic dependency, any communicable diseases and any chronic diseases (See Supplemental files).

### Statistical Analysis

We performed the univariate and bivariate analysis with suitable background characteristics. We have calculated absolute gaps in the prevalence of current own-perception and change in health status by background characteristics. The absolute gender gaps are in two folds defined as:

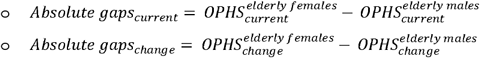

The study has then carried out an ordered logistic regression model (or proportional odds model) to determine the predictors of health perception among the elderly. The ordered logistic model is a regression model for an ordinal response variable (Grilli & Rampichini, 2014). Since the study has outcome variables with more than two categories and the values of each category have a sequential order and meet the criteria of proportional odds assumption. The study has used an ordered logistic regression model in terms of proportional odds ratios. The proportional odds ratios are estimated by exponentiating the ordered logit coefficients (UCLA: Statistical Consulting, 2021).

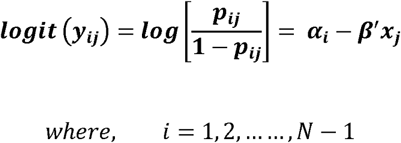

The parameter α_i_ is known as cut points (or threshold). They are in increasing orders here, are as follow:

1. poor<good<excellent
2. Worst<nearly same<better than before

## Results

### Sample profile

Table 1 shows the sample profile by gender with suitable socio-economic, demographic, and health characteristics among the elderly in India from the period (2017-18). There are 65% young-old women & 64% young-old and oldest woman (9%) are higher than most aged men (8%) while middle-old women (25%) are lower than middle-old men (27%). The majority of both older women & men belonged to the rural residence, Southern region, Hindu religion, most affluent group. About 63% of older women & 35% of older men have no education. Older women have marginally lower insurance coverage than men. There are 68% older women who are entirely economically dependent, whereas older men are only 27%. About 88% of older women & 86% of older men live with others other than their spouse. However, chronic diseases are higher among older women than men.

**Table 1.** Sample distribution of health perception among the elderly in India by gender with suitable background characteristics, 2017-18. (n=42,759).

| Background | Men |  | Women |  |
| --- | --- | --- | --- | --- |
|  | Samples | Percentage (%) | Samples | Percentage (%) |
| **Age-groups** |  |  |  |  |
| Young old | 14,094 | 64.35 | 13,674 | 65.56 |
| Middle old | 5,977 | 27.29 | 5,256 | 25.2 |
| Oldest old | 1,831 | 8.36 | 1,927 | 9.24 |
| **Place of residence** |  |  |  |  |
| Rural | 12,108 | 55.28 | 11,489 | 55.08 |
| Urban | 9,794 | 44.72 | 9,368 | 44.92 |
| **Regions** |  |  |  |  |
| Northern | 4,454 | 20.34 | 4,340 | 20.81 |
| Nort-Eastern | 2,169 | 9.9 | 1,820 | 8.73 |
| Central | 3,256 | 14.87 | 3,082 | 14.78 |
| Eastern | 3,672 | 16.77 | 3,314 | 15.89 |
| Western | 3,076 | 14.04 | 3,110 | 14.91 |
| Southern | 5,275 | 24.08 | 5,191 | 24.89 |
| **Marital status** |  |  |  |  |
| Never* | 3,392 | 15.49 | 10,045 | 48.16 |
| married | 18,510 | 84.51 | 10,812 | 51.84 |
| **Education status** |  |  |  |  |
| No-education | 7,746 | 35.37 | 13,123 | 62.92 |
| primary | 7,238 | 33.05 | 5,145 | 24.67 |
| secondary | 4,429 | 20.22 | 1,699 | 8.15 |
| Higher | 2,489 | 11.36 | 890 | 4.27 |
| **Religion** |  |  |  |  |
| Hindu | 16,979 | 77.52 | 16,261 | 77.96 |
| Non-hindu | 4,923 | 22.48 | 4,596 | 22.04 |
| **Caste groups** |  |  |  |  |
| SC/ST | 5,153 | 23.53 | 4,891 | 23.45 |
| OBC | 8,416 | 38.43 | 8,103 | 38.85 |
| General | 8,333 | 38.05 | 7,863 | 37.7 |
| **Wealth index** |  |  |  |  |
| Poorest | 3,666 | 16.74 | 3,525 | 16.9 |
| Poorer | 3,622 | 16.54 | 3,525 | 16.9 |
| Middle | 4,153 | 18.96 | 3,975 | 19.06 |
| Richer | 4,960 | 22.65 | 4,673 | 22.4 |
| Richest | 5,501 | 25.12 | 5,159 | 24.74 |
| **HH-members** |  |  |  |  |
| <= 5 | 10,556 | 48.2 | 10,644 | 51.03 |
| >5 | 11,346 | 51.8 | 10,213 | 48.97 |
| **HH Type** |  |  |  |  |
| Self employed | 13,900 | 63.46 | 13,007 | 62.36 |
| Regular wages | 2,107 | 9.62 | 2,237 | 10.73 |
| Casual workers | 2,469 | 11.27 | 2,508 | 12.02 |
| Others | 3,426 | 15.64 | 3,105 | 14.89 |
| **Insurance status** |  |  |  |  |
| Uncovered | 17,286 | 78.92 | 16,599 | 79.58 |
| Covered | 4,616 | 21.08 | 4,258 | 20.42 |
| **Economically** |  |  |  |  |
| **independence** |  |  |  |  |
| Independent | 10,598 | 48.39 | 1,812 | 8.69 |
| Partial | 5,328 | 24.33 | 4,801 | 23.02 |
| Fully | 5,976 | 27.29 | 14,244 | 68.29 |
| **Living arrangements** |  |  |  |  |
| Alone | 208 | 0.95 | 633 | 3.03 |
| Only with spouse | 2,791 | 12.74 | 1,758 | 8.43 |
| Others* | 18,903 | 86.31 | 18,466 | 88.54 |
| **Place of stay** |  |  |  |  |
| Other's house | 1,286 | 5.87 | 2,724 | 13.06 |
| Owned house | 20,616 | 94.13 | 18,133 | 86.94 |
| **Any communicable diseases** |  |  |  |  |
| No | 21,401 | 97.71 | 20,388 | 97.75 |
| Yes | 501 | 2.29 | 469 | 2.25 |
| **Any chronic diseases** |  |  |  |  |
| No | 16,817 | 76.78 | 15,846 | 75.97 |
| Yes | 5,085 | 23.22 | 5,011 | 24.03 |
| **Total** | **21,902** | **100** | **20,857** | **100** |

**Source:** Authors' own calculation using 75th round of National Sample Survey data.
**Note:** Never\*: includes elderly who were never married or separated or divorced. Non-Hindu\*: includes elderly who were belong to Muslims or Christians or Sikhs or Jains or Buddhists or others. Alone\*: As an inmate of old age homes or not as an inmate of old age homes. Others\*: without spouse but with children or other relations or non-relations. **Abbreviations:** SC- Schedule Caste; ST-Schedule Tribe; OBC- Other Backward Caste.

### Gender gaps in own-perception about current health status

Table 2 shows absolute gender gaps (%) in own-perception about current health status among the elderly in India from 2017-18. About 4% absolute gender gaps (AGG) are observed in poor OPHS_current_ among both young-old and middle-old age groups, which are higher than the oldest-old age. AGG in poor OPHS_current_ is higher in rural (4.7%) than urban areas (3.4%). North-eastern regions (7.65) have the highest AGG in poor OPHS_current_, whereas Southern regions have the lowest (3.4%). Marginally higher AGG in poor OPHS_current_ is seen among non-married than married and more than five-HH-members. No education (3.6%), Non-Hindu (8.6%), OBC (4.5%), SC/ST (4.3%), and Poorest (8.8%) have the highest AGG in poor OPHS_current_. Despite that, uncovered insurance support (4.6%) has greater AGG in poor OPHS_current_ than covered insurance (2.6%). Higher AGG in poor OPHS_current_ is reflected among those who are not dependent economically (5.5%), live with others other than a spouse (4.4%), and owned-house (4%). However, lower AGG in poor OPHS_current_ is found among the elderly with communicable diseases (2.5%) but more significant among those with chronic diseases (5%).

**Table 2.**
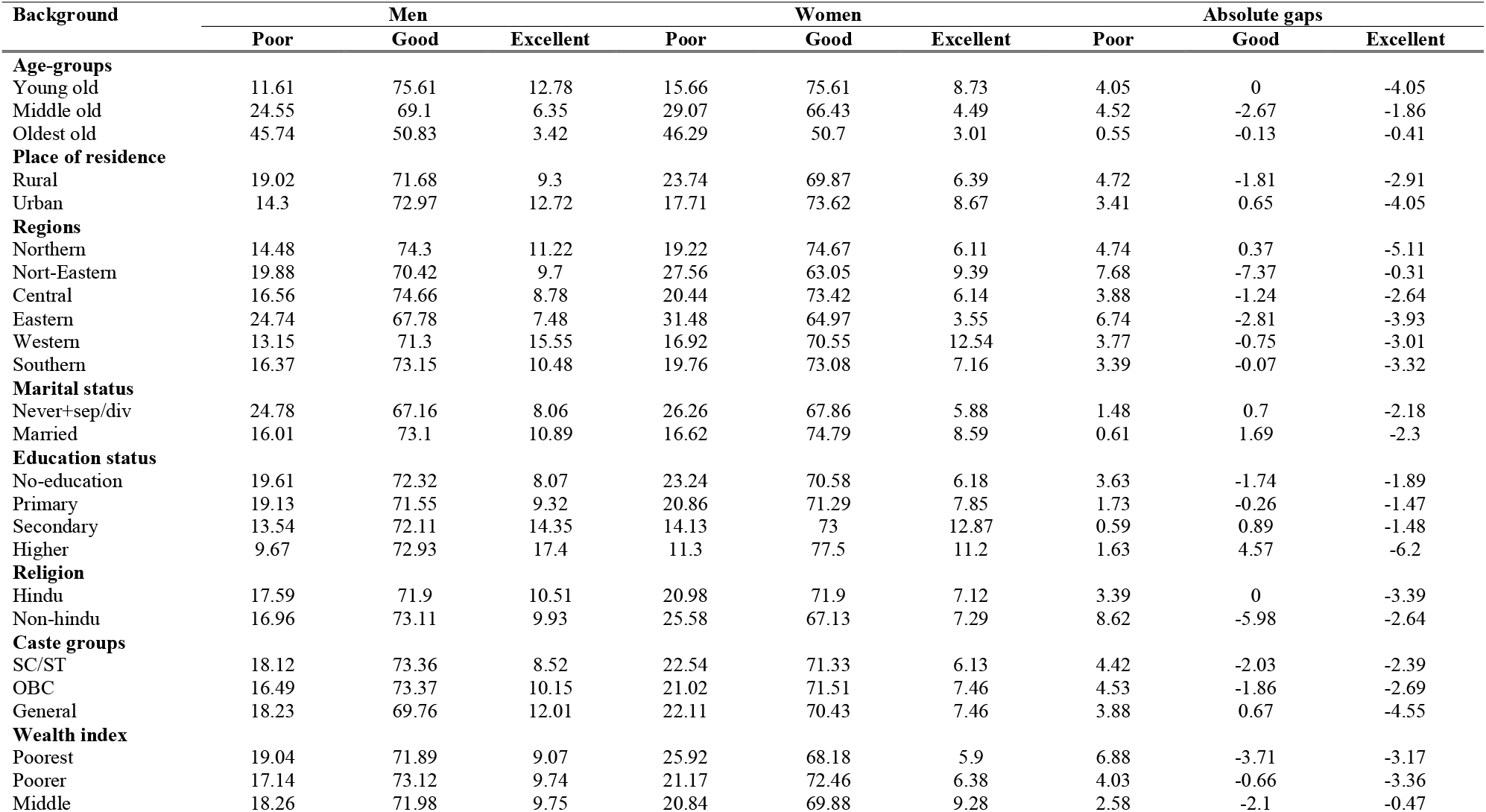
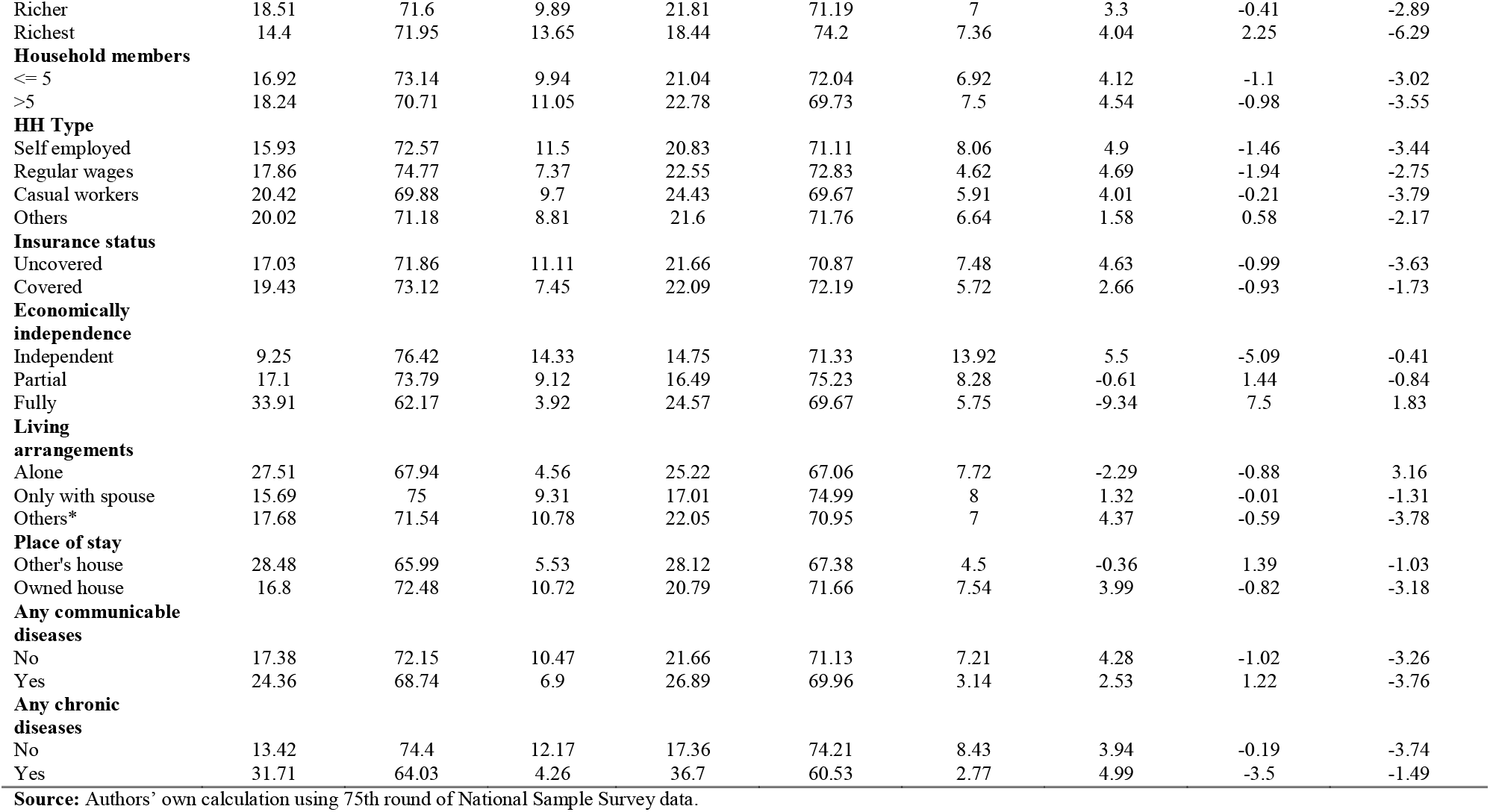
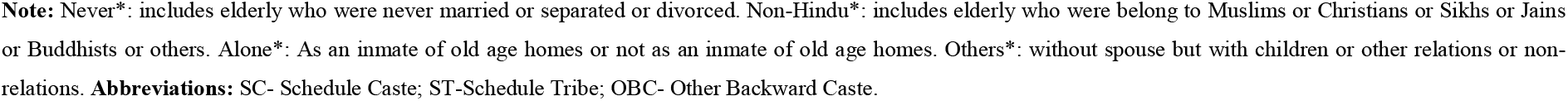
Absolute gender gaps (%) in own-perception about current health status among the elderly in India by gender with suitable background characteristics, 2017-18 (n=42,759).

| Background | Men |  |  | Women |  |  | Absolute gaps |  |  |
| --- | --- | --- | --- | --- | --- | --- | --- | --- | --- |
|  | Poor | Good | Excellent | Poor | Good | Excellent | Poor | Good | Excellent |
| **Age-groups** |  |  |  |  |  |  |  |  |  |
| Young old | 11.61 | 75.61 | 12.78 | 15.66 | 75.61 | 8.73 | 4.05 | 0 | -4.05 |
| Middle old | 24.55 | 69.1 | 6.35 | 29.07 | 66.43 | 4.49 | 4.52 | -2.67 | -1.86 |
| Oldest old | 45.74 | 50.83 | 3.42 | 46.29 | 50.7 | 3.01 | 0.55 | -0.13 | -0.41 |
| **Place of residence** |  |  |  |  |  |  |  |  |  |
| Rural | 19.02 | 71.68 | 9.3 | 23.74 | 69.87 | 6.39 | 4.72 | -1.81 | -2.91 |
| Urban | 14.3 | 72.97 | 12.72 | 17.71 | 73.62 | 8.67 | 3.41 | 0.65 | -4.05 |
| **Regions** |  |  |  |  |  |  |  |  |  |
| Northern | 14.48 | 74.3 | 11.22 | 19.22 | 74.67 | 6.11 | 4.74 | 0.37 | -5.11 |
| Nort-Eastern | 19.88 | 70.42 | 9.7 | 27.56 | 63.05 | 9.39 | 7.68 | -7.37 | -0.31 |
| Central | 16.56 | 74.66 | 8.78 | 20.44 | 73.42 | 6.14 | 3.88 | -1.24 | -2.64 |
| Eastern | 24.74 | 67.78 | 7.48 | 31.48 | 64.97 | 3.55 | 6.74 | -2.81 | -3.93 |
| Western | 13.15 | 71.3 | 15.55 | 16.92 | 70.55 | 12.54 | 3.77 | -0.75 | -3.01 |
| Southern | 16.37 | 73.15 | 10.48 | 19.76 | 73.08 | 7.16 | 3.39 | -0.07 | -3.32 |
| **Marital status** |  |  |  |  |  |  |  |  |  |
| Never+sep/div | 24.78 | 67.16 | 8.06 | 26.26 | 67.86 | 5.88 | 1.48 | 0.7 | -2.18 |
| Married | 16.01 | 73.1 | 10.89 | 16.62 | 74.79 | 8.59 | 0.61 | 1.69 | -2.3 |
| **Education status** |  |  |  |  |  |  |  |  |  |
| No-education | 19.61 | 72.32 | 8.07 | 23.24 | 70.58 | 6.18 | 3.63 | -1.74 | -1.89 |
| Primary | 19.13 | 71.55 | 9.32 | 20.86 | 71.29 | 7.85 | 1.73 | -0.26 | -1.47 |
| Secondary | 13.54 | 72.11 | 14.35 | 14.13 | 73 | 12.87 | 0.59 | 0.89 | -1.48 |
| Higher | 9.67 | 72.93 | 17.4 | 11.3 | 77.5 | 11.2 | 1.63 | 4.57 | -6.2 |
| **Religion** |  |  |  |  |  |  |  |  |  |
| Hindu | 17.59 | 71.9 | 10.51 | 20.98 | 71.9 | 7.12 | 3.39 | 0 | -3.39 |
| Non-hindu | 16.96 | 73.11 | 9.93 | 25.58 | 67.13 | 7.29 | 8.62 | -5.98 | -2.64 |
| **Caste groups** |  |  |  |  |  |  |  |  |  |
| SC/ST | 18.12 | 73.36 | 8.52 | 22.54 | 71.33 | 6.13 | 4.42 | -2.03 | -2.39 |
| OBC | 16.49 | 73.37 | 10.15 | 21.02 | 71.51 | 7.46 | 4.53 | -1.86 | -2.69 |
| General | 18.23 | 69.76 | 12.01 | 22.11 | 70.43 | 7.46 | 3.88 | 0.67 | -4.55 |
| **Wealth index** |  |  |  |  |  |  |  |  |  |
| Poorrest | 19.04 | 71.89 | 9.07 | 25.92 | 68.18 | 5.9 | 6.88 | -3.71 | -3.17 |
| Poorer | 17.14 | 73.12 | 9.74 | 21.17 | 72.46 | 6.38 | 4.03 | -0.66 | -3.36 |
| Middle | 18.26 | 71.98 | 9.75 | 20.84 | 69.88 | 9.28 | 2.58 | -2.1 | -0.47 |
| Richer | 18.51 | 71.6 | 9.89 | 21.81 | 71.19 | 7 | 3.3 | -0.41 | -2.89 |
| Richest | 14.4 | 71.95 | 13.65 | 18.44 | 74.2 | 7.36 | 4.04 | 2.25 | -6.29 |
| **Household members** |  |  |  |  |  |  |  |  |  |
| <= 5 | 16.92 | 73.14 | 9.94 | 21.04 | 72.04 | 6.92 | 4.12 | -1.1 | -3.02 |
| >5 | 18.24 | 70.71 | 11.05 | 22.78 | 69.73 | 7.5 | 4.54 | -0.98 | -3.55 |
| **HH Type** |  |  |  |  |  |  |  |  |  |
| Self employed | 15.93 | 72.57 | 11.5 | 20.83 | 71.11 | 8.06 | 4.9 | -1.46 | -3.44 |
| Regular wages | 17.86 | 74.77 | 7.37 | 22.55 | 72.83 | 4.62 | 4.69 | -1.94 | -2.75 |
| Casual workers | 20.42 | 69.88 | 9.7 | 24.43 | 69.67 | 5.91 | 4.01 | -0.21 | -3.79 |
| Others | 20.02 | 71.18 | 8.81 | 21.6 | 71.76 | 6.64 | 1.58 | 0.58 | -2.17 |
| **Insurance status** |  |  |  |  |  |  |  |  |  |
| Uncovered | 17.03 | 71.86 | 11.11 | 21.66 | 70.87 | 7.48 | 4.63 | -0.99 | -3.63 |
| Covered | 19.43 | 73.12 | 7.45 | 22.09 | 72.19 | 5.72 | 2.66 | -0.93 | -1.73 |
| **Economically independence** |  |  |  |  |  |  |  |  |  |
| Independent | 9.25 | 76.42 | 14.33 | 14.75 | 71.33 | 13.92 | 5.5 | -5.09 | -0.41 |
| Partial | 17.1 | 73.79 | 9.12 | 16.49 | 75.23 | 8.28 | -0.61 | 1.44 | -0.84 |
| Fully | 33.91 | 62.17 | 3.92 | 24.57 | 69.67 | 5.75 | -9.34 | 7.5 | 1.83 |
| **Living arrangements** |  |  |  |  |  |  |  |  |  |
| Alone | 27.51 | 67.94 | 4.56 | 25.22 | 67.06 | 7.72 | -2.29 | -0.88 | 3.16 |
| Only with spouse | 15.69 | 75 | 9.31 | 17.01 | 74.99 | 8 | 1.32 | -0.01 | -1.31 |
| Others* | 17.68 | 71.54 | 10.78 | 22.05 | 70.95 | 7 | 4.37 | -0.59 | -3.78 |
| **Place of stay** |  |  |  |  |  |  |  |  |  |
| Other's house | 28.48 | 65.99 | 5.53 | 28.12 | 67.38 | 4.5 | -0.36 | 1.39 | -1.03 |
| Owned house | 16.8 | 72.48 | 10.72 | 20.79 | 71.66 | 7.54 | 3.99 | -0.82 | -3.18 |
| **Any communicable diseases** |  |  |  |  |  |  |  |  |  |
| No | 17.38 | 72.15 | 10.47 | 21.66 | 71.13 | 7.21 | 4.28 | -1.02 | -3.26 |
| Yes | 24.36 | 68.74 | 6.9 | 26.89 | 69.96 | 3.14 | 2.53 | 1.22 | -3.76 |
| **Any chronic diseases** |  |  |  |  |  |  |  |  |  |
| No | 13.42 | 74.4 | 12.17 | 17.36 | 74.21 | 8.43 | 3.94 | -0.19 | -3.74 |
| Yes | 31.71 | 64.03 | 4.26 | 36.7 | 60.53 | 2.77 | 4.99 | -3.5 | -1.49 |

**Source:** Authors' own calculation using 75th round of National Sample Survey data.
**Note:** Never\*: includes elderly who were never married or separated or divorced. Non-Hindu\*: includes elderly who were belong to Muslims or Christians or Sikhs or Jains or Buddhists or others. Alone\*: As an inmate of old age homes or not as an inmate of old age homes. Others\*: without spouse but with children or other relations or non-relations. **Abbreviations:** SC- Schedule Caste; ST-Schedule Tribe; OBC- Other Backward Caste.

Besides that, greater AGG in good OPHS_current_ is reflected among those with higher education (4.6%), Richest group (2.25%), and fully economically dependent (7.5%).

### Gender gaps in own-perception about change in health status

Table 3 shows absolute gender gaps (%) in own-perception about change in health status among the elderly in India from 2017-18. Around 3.1% AGG in nearly-same OPHS_change_ are found among oldest-old which is greater than the young-old (2%) & middle-old (1.6%). AGG in the nearly-same OPHS_change_ is lower among urban (1.3%) than rural (2.6%). Eastern regions (6%) have the highest AGG in nearly-same OPHS_change_, followed by North-eastern region (4.6%), Southern region (2%) but lowest is seen in the Northern region (1.6%). Unmarried (5.2%), primary education (3.7%), middle-income group (5.5%), covered insurance (4.3%), economically independent (6.45%), living alone (14.5%) & owned house (2.1%) have higher AGG in nearly-same OPHS_change_. Interestingly, Non-Hindus (1.7%) have marginally lower AGG in nearly-same OPHS_change_ than the Hindus (2.3%), but OBC (4.4%) has higher AGG in nearly-same OPHS_change_ than SC/ST (2.5%). Nearly 2.4% AGG in the nearly-same OPHS_change_ is seen among those with no communicable diseases.

**Table 3.**
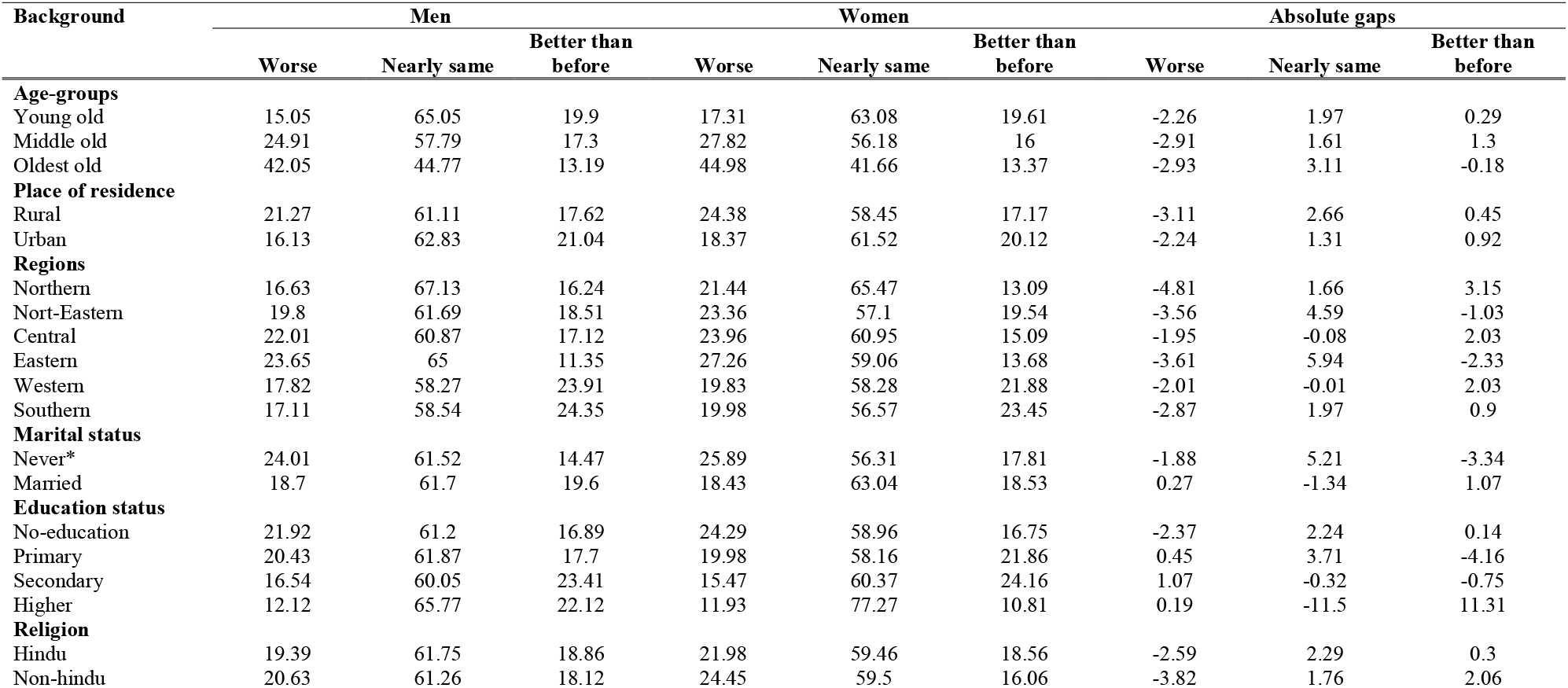
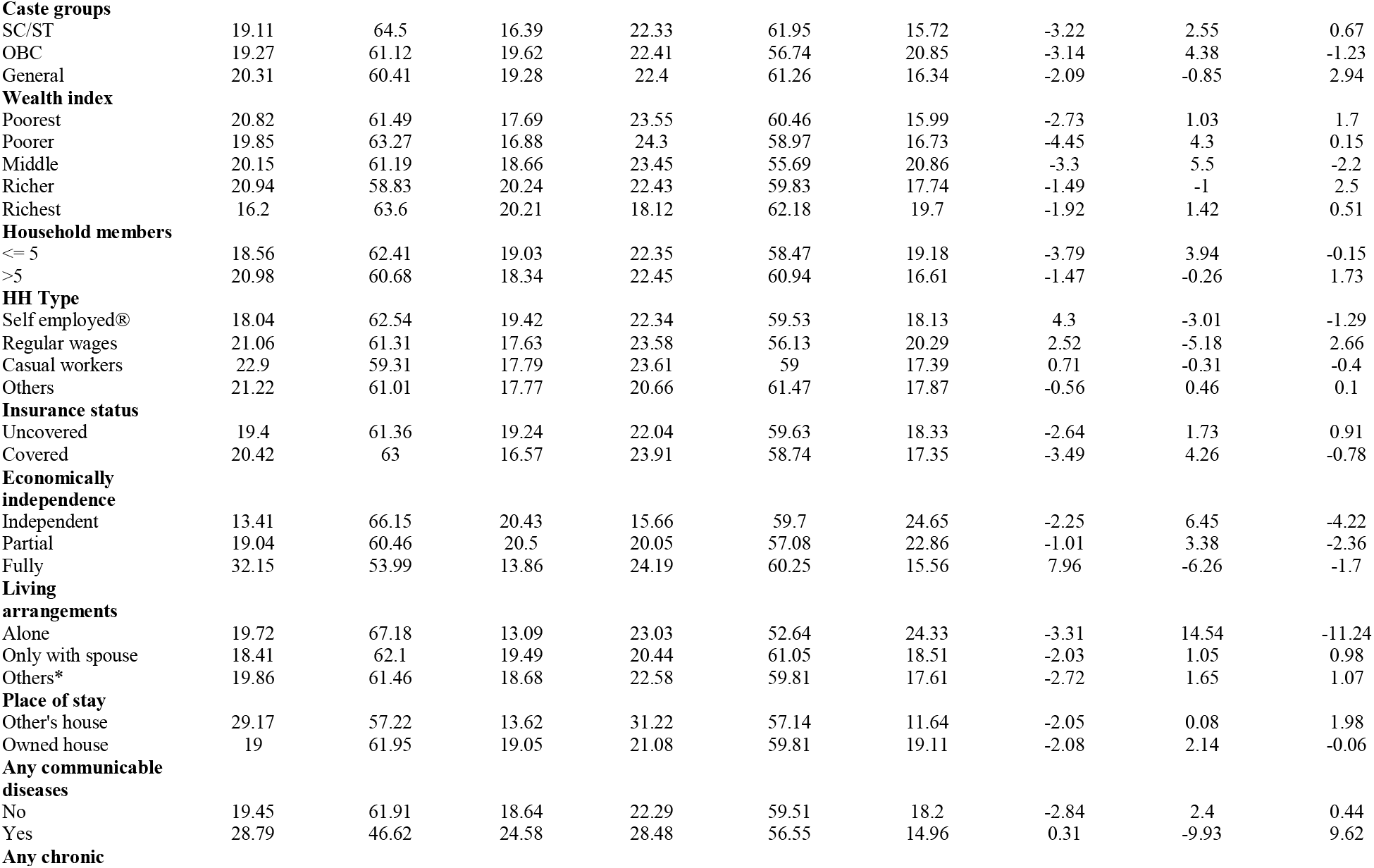
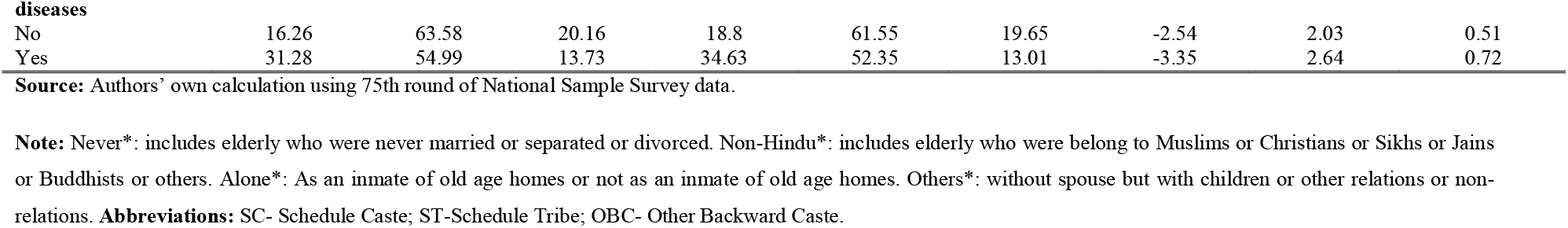
Absolute gender gaps (%) in own-perception about change in health status among the elderly in India by gender with suitable background characteristics, 2017-18. (n=42,759).

| Background | Men |  |  | Women |  |  | Absolute gaps |  |  |
| --- | --- | --- | --- | --- | --- | --- | --- | --- | --- |
|  | Worse | Nearly same | Better than before | Worse | Nearly same | Better than before | Worse | Nearly same | Better than before |
| **Age-groups** |  |  |  |  |  |  |  |  |  |
| Young old | 15.05 | 65.05 | 19.9 | 17.31 | 63.08 | 19.61 | -2.26 | 1.97 | 0.29 |
| Middle old | 24.91 | 57.79 | 17.3 | 27.82 | 56.18 | 16 | -2.91 | 1.61 | 1.3 |
| Oldest old | 42.05 | 44.77 | 13.19 | 44.98 | 41.66 | 13.37 | -2.93 | 3.11 | -0.18 |
| **Place of residence** |  |  |  |  |  |  |  |  |  |
| Rural | 21.27 | 61.11 | 17.62 | 24.38 | 58.45 | 17.17 | -3.11 | 2.66 | 0.45 |
| Urban | 16.13 | 62.83 | 21.04 | 18.37 | 61.52 | 20.12 | -2.24 | 1.31 | 0.92 |
| **Regions** |  |  |  |  |  |  |  |  |  |
| Northern | 16.63 | 67.13 | 16.24 | 21.44 | 65.47 | 13.09 | -4.81 | 1.66 | 3.15 |
| Nort-Eastern | 19.8 | 61.69 | 18.51 | 23.36 | 57.1 | 19.54 | -3.56 | 4.59 | -1.03 |
| Central | 22.01 | 60.87 | 17.12 | 23.96 | 60.95 | 15.09 | -1.95 | -0.08 | 2.03 |
| Eastern | 23.65 | 65 | 11.35 | 27.26 | 59.06 | 13.68 | -3.61 | 5.94 | -2.33 |
| Western | 17.82 | 58.27 | 23.91 | 19.83 | 58.28 | 21.88 | -2.01 | -0.01 | 2.03 |
| Southern | 17.11 | 58.54 | 24.35 | 19.98 | 56.57 | 23.45 | -2.87 | 1.97 | 0.9 |
| **Marital status** |  |  |  |  |  |  |  |  |  |
| Never* | 24.01 | 61.52 | 14.47 | 25.89 | 56.31 | 17.81 | -1.88 | 5.21 | -3.34 |
| Married | 18.7 | 61.7 | 19.6 | 18.43 | 63.04 | 18.53 | 0.27 | -1.34 | 1.07 |
| **Education status** |  |  |  |  |  |  |  |  |  |
| No-education | 21.92 | 61.2 | 16.89 | 24.29 | 58.96 | 16.75 | -2.37 | 2.24 | 0.14 |
| Primary | 20.43 | 61.87 | 17.7 | 19.98 | 58.16 | 21.86 | 0.45 | 3.71 | -4.16 |
| Secondary | 16.54 | 60.05 | 23.41 | 15.47 | 60.37 | 24.16 | 1.07 | -0.32 | -0.75 |
| Higher | 12.12 | 65.77 | 22.12 | 11.93 | 77.27 | 10.81 | 0.19 | -11.5 | 11.31 |
| **Religion** |  |  |  |  |  |  |  |  |  |
| Hindu | 19.39 | 61.75 | 18.86 | 21.98 | 59.46 | 18.56 | -2.59 | 2.29 | 0.3 |
| Non-hindu | 20.63 | 61.26 | 18.12 | 24.45 | 59.5 | 16.06 | -3.82 | 1.76 | 2.06 |
| Caste groups |  |  |  |  |  |  |  |  |  |
| SC/ST | 19.11 | 64.5 | 16.39 | 22.33 | 61.95 | 15.72 | -3.22 | 2.55 | 0.67 |
| OBC | 19.27 | 61.12 | 19.62 | 22.41 | 56.74 | 20.85 | -3.14 | 4.38 | -1.23 |
| General | 20.31 | 60.41 | 19.28 | 22.4 | 61.26 | 16.34 | -2.09 | -0.85 | 2.94 |
| Wealth index |  |  |  |  |  |  |  |  |  |
| Poorest | 20.82 | 61.49 | 17.69 | 23.55 | 60.46 | 15.99 | -2.73 | 1.03 | 1.7 |
| Poorer | 19.85 | 63.27 | 16.88 | 24.3 | 58.97 | 16.73 | -4.45 | 4.3 | 0.15 |
| Middle | 20.15 | 61.19 | 18.66 | 23.45 | 55.69 | 20.86 | -3.3 | 5.5 | -2.2 |
| Richer | 20.94 | 58.83 | 20.24 | 22.43 | 59.83 | 17.74 | -1.49 | -1 | 2.5 |
| Richest | 16.2 | 63.6 | 20.21 | 18.12 | 62.18 | 19.7 | -1.92 | 1.42 | 0.51 |
| Household members |  |  |  |  |  |  |  |  |  |
| <= 5 | 18.56 | 62.41 | 19.03 | 22.35 | 58.47 | 19.18 | -3.79 | 3.94 | -0.15 |
| >5 | 20.98 | 60.68 | 18.34 | 22.45 | 60.94 | 16.61 | -1.47 | -0.26 | 1.73 |
| HH Type |  |  |  |  |  |  |  |  |  |
| Self employed® | 18.04 | 62.54 | 19.42 | 22.34 | 59.53 | 18.13 | 4.3 | -3.01 | -1.29 |
| Regular wages | 21.06 | 61.31 | 17.63 | 23.58 | 56.13 | 20.29 | 2.52 | -5.18 | 2.66 |
| Casual workers | 22.9 | 59.31 | 17.79 | 23.61 | 59 | 17.39 | 0.71 | -0.31 | -0.4 |
| Others | 21.22 | 61.01 | 17.77 | 20.66 | 61.47 | 17.87 | -0.56 | 0.46 | 0.1 |
| Insurance status |  |  |  |  |  |  |  |  |  |
| Uncovered | 19.4 | 61.36 | 19.24 | 22.04 | 59.63 | 18.33 | -2.64 | 1.73 | 0.91 |
| Covered | 20.42 | 63 | 16.57 | 23.91 | 58.74 | 17.35 | -3.49 | 4.26 | -0.78 |
| Economically independence |  |  |  |  |  |  |  |  |  |
| Independent | 13.41 | 66.15 | 20.43 | 15.66 | 59.7 | 24.65 | -2.25 | 6.45 | -4.22 |
| Partial | 19.04 | 60.46 | 20.5 | 20.05 | 57.08 | 22.86 | -1.01 | 3.38 | -2.36 |
| Fully | 32.15 | 53.99 | 13.86 | 24.19 | 60.25 | 15.56 | 7.96 | -6.26 | -1.7 |
| Living arrangements |  |  |  |  |  |  |  |  |  |
| Alone | 19.72 | 67.18 | 13.09 | 23.03 | 52.64 | 24.33 | -3.31 | 14.54 | -11.24 |
| Only with spouse | 18.41 | 62.1 | 19.49 | 20.44 | 61.05 | 18.51 | -2.03 | 1.05 | 0.98 |
| Others* | 19.86 | 61.46 | 18.68 | 22.58 | 59.81 | 17.61 | -2.72 | 1.65 | 1.07 |
| Place of stay |  |  |  |  |  |  |  |  |  |
| Other's house | 29.17 | 57.22 | 13.62 | 31.22 | 57.14 | 11.64 | -2.05 | 0.08 | 1.98 |
| Owned house | 19 | 61.95 | 19.05 | 21.08 | 59.81 | 19.11 | -2.08 | 2.14 | -0.06 |
| Any communicable diseases |  |  |  |  |  |  |  |  |  |
| No | 19.45 | 61.91 | 18.64 | 22.29 | 59.51 | 18.2 | -2.84 | 2.4 | 0.44 |
| Yes | 28.79 | 46.62 | 24.58 | 28.48 | 56.55 | 14.96 | 0.31 | -9.93 | 9.62 |
| Any chronic |  |  |  |  |  |  |  |  |  |
| **diseases** |  |  |  |  |  |  |  |  |  |
| No | 16.26 | 63.58 | 20.16 | 18.8 | 61.55 | 19.65 | -2.54 | 2.03 | 0.51 |
| Yes | 31.28 | 54.99 | 13.73 | 34.63 | 52.35 | 13.01 | -3.35 | 2.64 | 0.72 |

**Source:** Authors' own calculation using 75th round of National Sample Survey data.
**Note:** Never\*: includes elderly who were never married or separated or divorced. Non-Hindu\*: includes elderly who were belong to Muslims or Christians or Sikhs or Jains or Buddhists or others. Alone\*: As an inmate of old age homes or not as an inmate of old age homes. Others\*: without spouse but with children or other relations or non-relations. **Abbreviations:** SC- Schedule Caste; ST-Schedule Tribe; OBC- Other Backward Caste.

However, young-old age has lower AGG (0.3%) in much-better-OPHS_change_ than middle-old age (1.3%). Urban areas have marginally higher AGG in much-better-OPHS_change_ than rural. Highest AGG in much-better-OPHS_change_ is found in the Northern region (3.15%), followed by the Eastern region (2.3%), Central region (2%), Western region (2%), and lowest in the Southern region (0.9%). Only 1% AGG in much-better-OPHS_change_ is seen among married, fully mobile, and uncovered insurance. Negligible AGG in much-better-OPHS_change_ is observed among those with no education (0.1%) but significantly higher among Higher education (11.3%). Non-Hindu religion (2.06%), General caste (3%), and Richer group (2.5%) have the highest AGG in much-better-OPHS_change_. More than five HH-members are facing greater AGG in much-better-OPHS_change_. Interestingly, significantly greater AGG in much-better-OPHS_change_ is observed among the elderly with communicable diseases (10%). Still, there is marginal AGG in much-better-OPHS_change_ reflected among those with chronic diseases.

### Determinants of (OPHS_current_) and (OPHS_change_)

Table 4 shows the result of ordered regression analysis of own-perception about the current (OPHS_current_) & change (OPHS_change_) in health status among the elderly in India with suitable background characteristics, 2017-18. **Model 1** in **Table 4** shows that odds of being rated good or excellent (versus poor) are found to be significantly higher among women [OR=1.27; CI=1.21, 1.34] than men. The middle-old [OR=0.5; CI=0.47, 0.52] and oldest-old [OR=0.27; CI=0.25, 0.29] have significantly lower odds of being rated good or excellent (versus poor) compared to young old. The elderly belonging to the Western [OR=1.67; CI=1.55, 1.8] and Southern [OR=1.3; CI=1.21, 1.39] regions had significantly greater odds of being rated good or excellent (versus poor) compared to Northern regions while the North-Eastern, Central and Eastern regions showed lower. The odds ratio of being rated good or excellent (versus poor) is also greater and significant for socio-economic factors like marital status, education, and income (only richer and richest group). The results indicate the greater preference odds of health perception among the household member more than five [OR=1.12; CI=1.07, 1.18] compared to household member &#x2264;5. Lower odds of being rated good or excellent (versus poor) were found among the elderly who receives regular wages, casual wages, and others compared to self-employed.

**Table 4.**
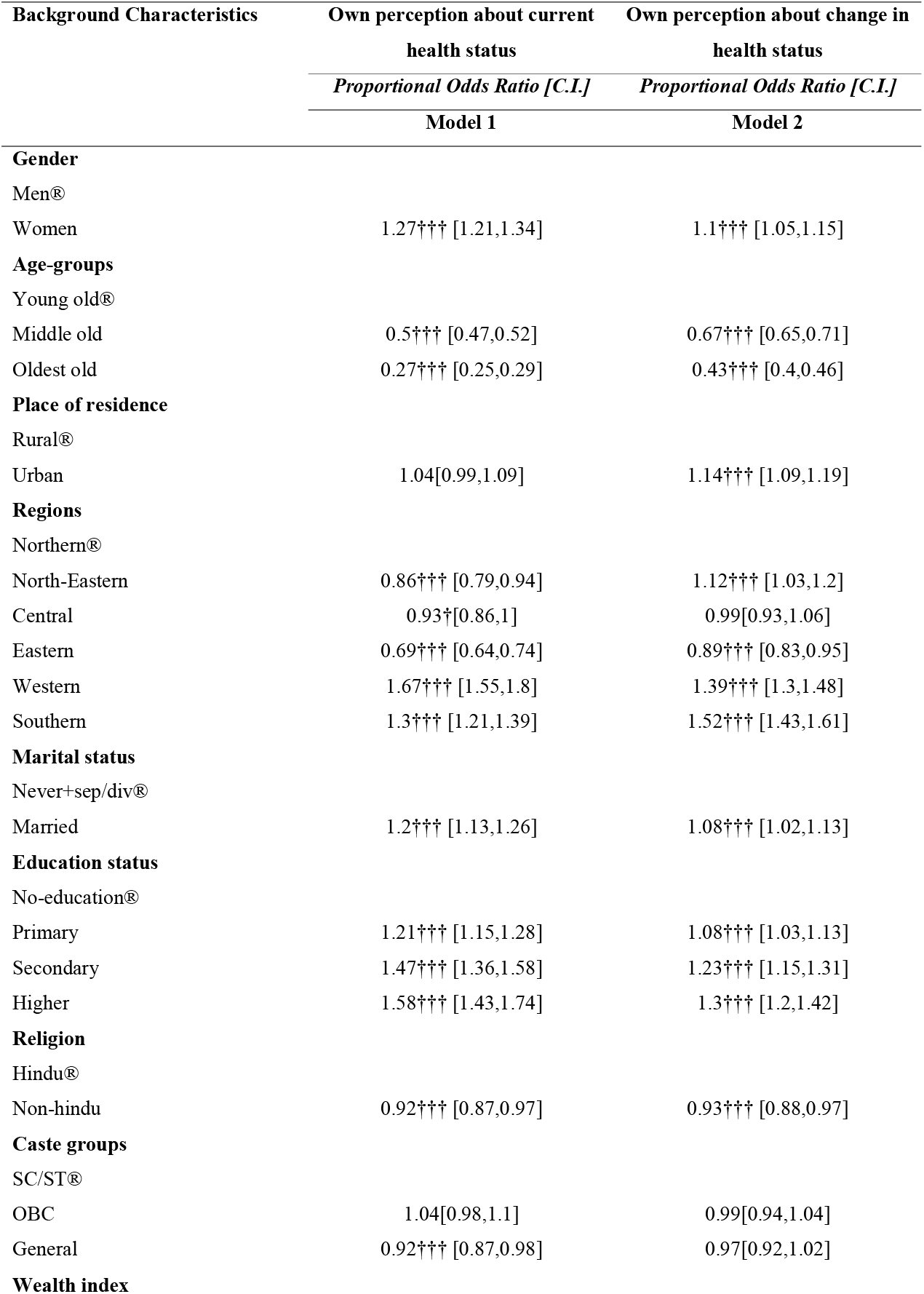
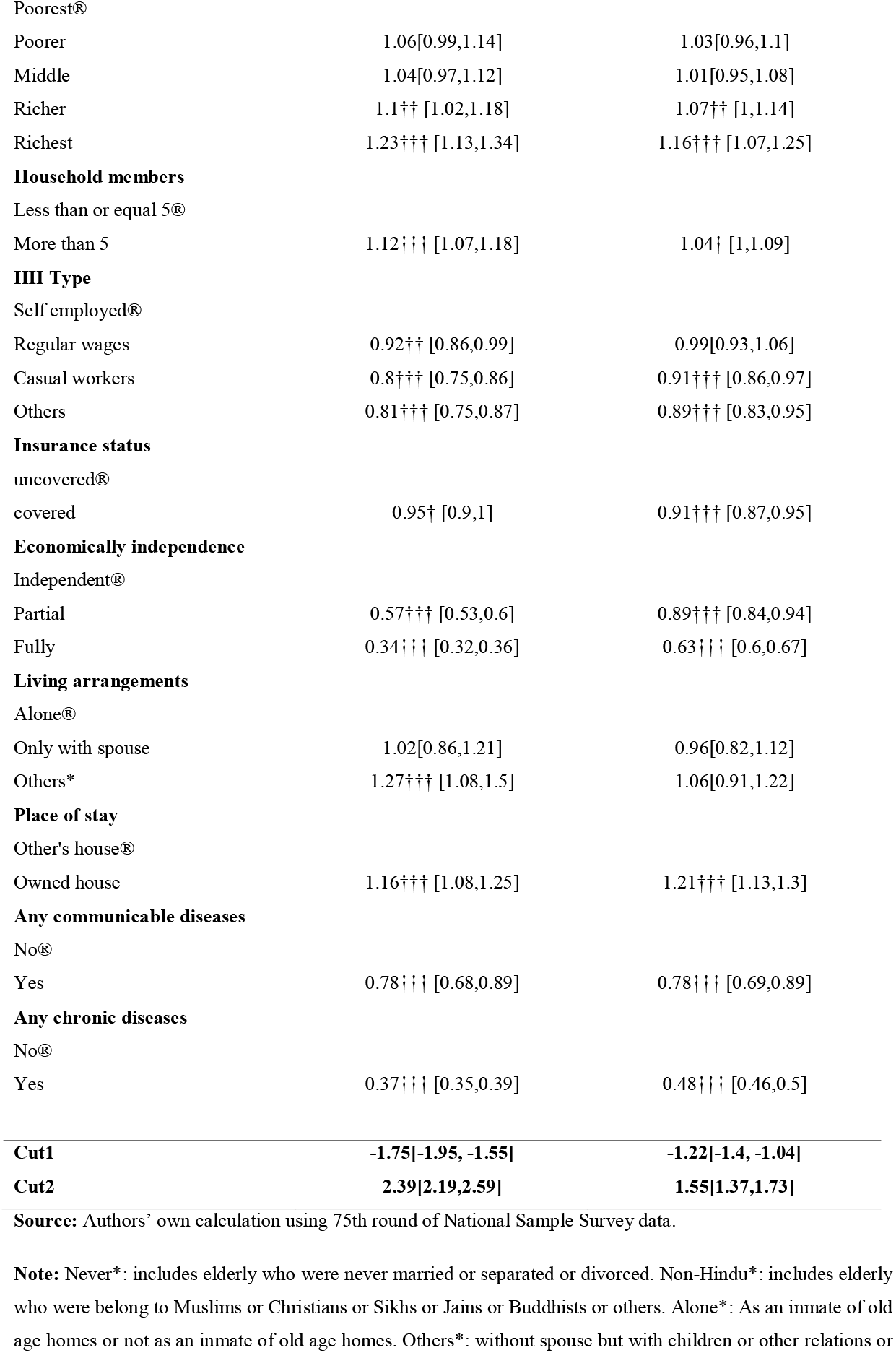
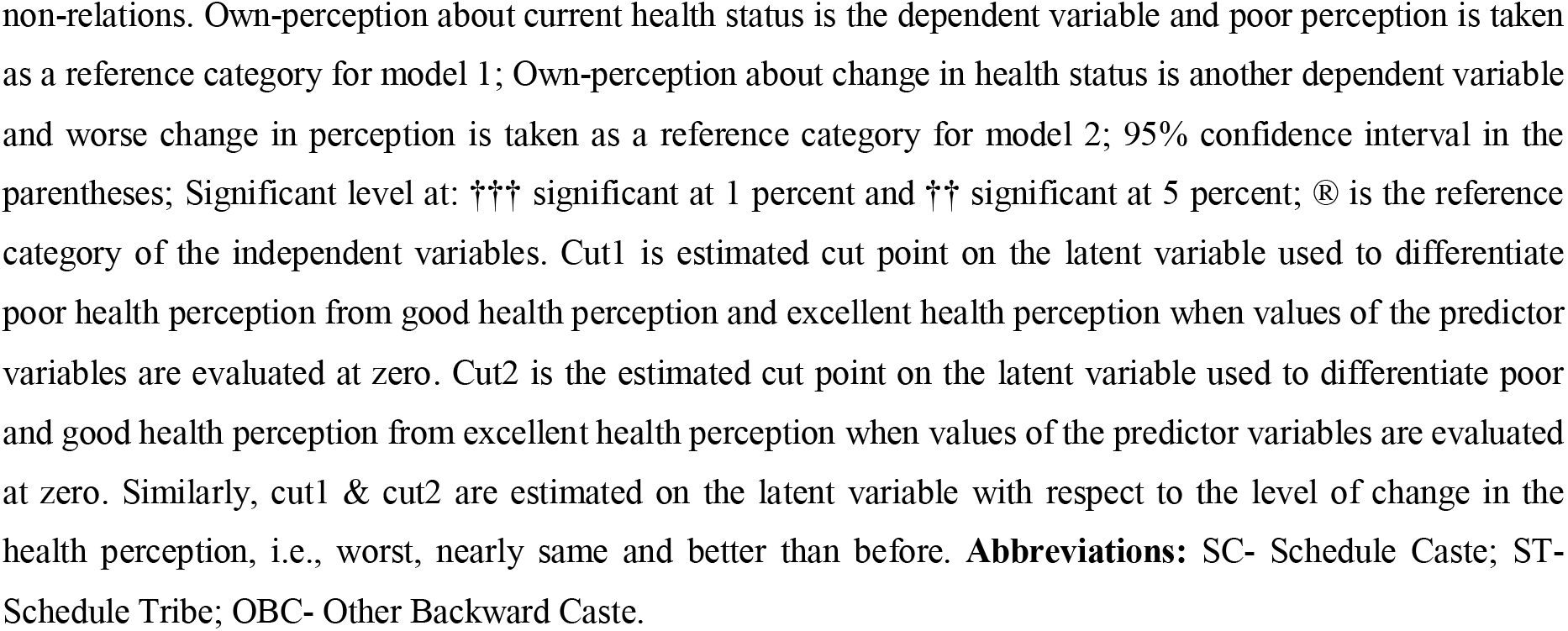
Ordered regression analysis of own-perception about current & change in health status among the elderly in India with suitable background characteristics, 2017-18. (n=42,759).

Despite that, we have also examined key risk predictors like economic independence, living arrangements, place of stay, chronic diseases, communicable diseases and health insurance, and family size. The elderly covered with health insurance, partial and fully economically dependent, were at lower odds. In contrast, elderly living with others [OR=1.27; CI=1.08, 1.5] show greater odds of having a good or excellent perception of health than living alone. At the same time, it has been found that elderly who stay at their own house [OR=1.16; CI=1.08, 1.25] show higher odds of having a good or excellent perception of health than staying in others’ houses. The odds of being rated good or excellent perception about health (versus poor) is found lower with any communicable disease conditions, as can be seen from the odds ratios [OR=0.78; CI=0.68, 0.89], and similar results is being found with any chronic diseases [OR=0.37; CI=0.35, 0.39] respectively.

However, **Model 2** in **Table 4** shows the results of ordered logistic regression for own perception about the change in health status. The model’s outcome variable was categorized as (nearly-same or better than before (versus worse) with worse change as the reference category. The result shows a clear and significant association between change in health perception and socio-economic and other risk factors. Socio-economic factors like gender, residence, wealth quintile and household size have positive and significant associations. The odds of being rated nearly-same or better (versus worse) change in health perception were found to be higher among older women [OR=1.1; CI=1.05, 1.15] compared to older men. While the lower odds of a change in health perception were found among middle-old and oldest-old compared to young-old, similar results were found in OPHScurrent. Now, compared to the rural residence, urban residence [OR=1.14; CI=1.09, 1.19] is showing higher odds of being rated same or better (versus worse) change in health perception. Southern, Western and Northeastern regions show greater while the eastern region showed lower odds of a change in health perception than the Northern region. The socio-economic factors like marital status, educational status, and income (only richer and richest group) reflect the higher odds of being rated the same or better (versus worse) change in health perception. The greater odds of being rated nearly-same or better (versus worse) change in health perception was found among the household member more than five [OR=1.04; CI=1, 1.09] than household member &#x2264;5. Lower odds of being rated nearly-same or better (versus worse) were found among the elderly who receive only casual wages and others than self-employed.

Notwithstanding, the insured elderly, partially and fully economically dependent elderly have lower odds of being rated same or better (versus worse) change in health perception, while the elderly who stays at their own house exhibited higher. Elderly with communicable diseases [OR=0.78; CI=0.69, 0.89], and chronic diseases [OR=048; CI=0.46, 0.5] showing lower odds of being rated same or better (versus worse) change in health perception.

## Discussion

We have used India’s large-scale national sample survey data, where we have examined not only the current health perception but also analyzed it to study the change in health perception among the elderly from a gender perspective. The perception about the health status of the reflects the health condition which is a significant predictor for active and healthy aging. Given the rapid growth of the elderly and increasing life longevity, quality of life at upper ages are the significant factors to be studied from a public health perspective. While many factors determine the quality of life at older ages, one of the significant factors is the own perception of the elderly, which they want to perceive about their health (Bhan et al., 2017). Therefore, we attempted to examine the current health perception and the change in health perception among the elderly in India, owing to the notion that perception about health is one of the significant factors that shapers the better quality of life at later ages (Akhtar & Saikia, 2021; Ranjan & Muraleedharan, 2020; Smith et al., 2012).

### Current health perception

Our results showed a clear and significant association between health perception and socio-economic outcomes. While there is a clear gap between men and women in terms of rating poor perception about health, men generally have reported higher perceptions about their current status of health when it comes to rating them excellent in terms of socio-economic outcomes like income, place of residence, and household structure or size. These results were similar to earlier findings where women generally report poor perceptions about their health stats (Welin et al., 2011). This signifies the importance of better socio-economic conditions from a gender perspective, where women with better socio-economic conditions likely indicate a lower perception of health (Rathbun et al., 2020).

An earlier study has suggested that health perception is affected by health behavior, and it is also attributed to illness, especially to the advanced aged because of the chronic disease, which is unavoidable support to an older age (Dohrmann, 2018). We have also examined the factors like morbidity in which both communicable diseases and chronic diseases showed lower odds of being rated good or excellent in health perception and our finding is also supported by the previous studies (Barreto & Figueiredo, 2009; Cramm & Nieboer, 2016; Diener & Chan, 2011; Dohrmann, 2018).

These factors determine the differences in gender perception of health status were similar to what we witnessed in socio-economic factors like income, residence, family size, and education, where a greater proportion of women reported poor health perception as compared to men (Krewski et al., 2006; Lemyre et al., 2006; Linder et al., 2010). These findings corroborate with earlier results in many other studies reflecting the asymmetry in gender dimension of health perception about their current health (Linder et al., 2010). Factors like economic dependency and health insurance are also key to health perception. The results from our study confirm this association. Similarly, results were found in terms of economic independence reflecting the fact that health perception is independent of whether being insured or financially independent by gender perspective (Ha & Kim, 2019).

### Change in health perception

The results in the study were slightly contrasting to what we have obtained about the perception of the current status of health. Although there is a significant and clear association of change in current perception of health with various risk factors as found in the earlier studies (Machón et al., 2016). The perception of women’s health has significantly similar in this case. The results are identical to what we witnessed in Model 1 (Table 4). The change in perception among women was found to be higher as compared to men both in terms of socio-economic and health outcomes (Forte et al., 2015). Our finding has also showed that the urban residence and married elderly have higher odds of being rated nearly-same or better change in health perception and similar results have also been found in the previous studies (Ellis et al., 2021; Goldstein et al., 1984).

However, the factors like educational status of the elderly, income status, and household size also reported better perception about change in health perception after hospitalization (De-Nour & Shanan, 1966). Previous studies have clearly reflected that the perception that can vary once the change in health status is improved among the women (Warmoth et al., 2016; Watcharanat et al., 2019).

The current study provides a detailed analysis of health perception and its gender perspective among the elderly in India. At the same time, the results confirm that women report more likely poor health as compared to men given various socio-economic and health outcomes. A previous study (Husain & Ghosh, 2017) has suggested that there is slightly a better perception about health when we measure the change in current perception of health among women as compared to men. Despite some limitations, this study tried to make a significant account of not only perception about the current health status but also change in the perception of current health status among the elderly.

### Limitations

The study could not include the various key factors while examining the health perception phenomena. Factors like body mass index, frailty, and other nutritional health outcomes could not be examined since the data was not available about them in the sample taken for consideration. Secondly, the study was mainly conducted on the elderly aged 50 and above, which might not be the case in terms of the younger population given their less vulnerability to health and socio-economic risks. Moreover, there is already a challenge of age-related discrimination and greater disability likelihood at upper ages, which might have significantly affected the results in this study.

## Conclusions

Despite numerous limitations, this study addressed the significant public health concern, which is key to addressing the challenge of the elderly health and their perception of well-being. The elderly are more vulnerable to health and physical outcomes given the age-related life cycle changes, so the increased risk for active and healthy aging is likely a challenge given the low perception about current health status. Moreover, the challenges are multiple given the asymmetry from a gender perspective since women are more prone to these health outcomes, which likely risks their well-being. Therefore, this study identifies a significant gender gap in this domain since identifying the elderly health perception can be significant in terms of their healthcare services and caregiving approaches.

## Data Availability

The data can be downloadable using the link: https://www.mospi.gov.in/web/mospi/download-tables-data/-/reports/view/templateTwo/16202?q=TBDCAT

## Acknowledgement

**None**

## Reference

Akhtar, S. N., & Saikia, N. (2021). Differentials and predictors of hospitalization among the elderly people in India: Evidence from 75th round of National Sample Survey (2017−18) [Preprint]. Public and Global Health. https://doi.org/10.1101/2021.08.25.21262606

Barreto, S. M., & Figueiredo, R.C. de. (2009). Doença crônica, auto-avaliação de saúde e comportamento de risco: Diferença de gênero. Revista de Saúde Pública, 43(Suppl 2), 38–47. https://doi.org/10.1590/S0034-89102009000900006

Bhan, N., Madhira, P., Muralidharan, A., Kulkarni, B., Murthy, G., Basu, S., & Kinra, S. (2017). Health needs, access to healthcare, and perceptions of ageing in an urbanizing community in India: A qualitative study. BMC Geriatrics, 17(1), 156. https://doi.org/10.1186/s12877-017-0544-y

Bora, J. K., & Saikia, N. (2015). Gender Differentials in Self-Rated Health and Self-Reported Disability among Adults in India. PLOS ONE, 10(11), e0141953. https://doi.org/10.1371/journal.pone.0141953

Cramm, J. M., & Nieboer, A. P. (2016). Is “disease management” the answer to our problems? No! Population health management and (disease) prevention require “management of overall well-being.” BMC Health Services Research, 16(1), 500. https://doi.org/10.1186/s12913-016-1765-z

De-Nour, A. K., & Shanan, J. (1966). Changes in Self Perception During Hospitalisation in an Open Ward of a General Hospital. Psychotherapy and Psychosomatics, 14(2), 133–143. https://www.jstor.org/stable/45112860?seq=1#metadata_info_tab_contents

Diener, E., & Chan, M. Y. (2011). Happy People Live Longer: Subjective Well-Being Contributes to Health and Longevity: HEALTH BENEFITS OF HAPPINESS. Applied Psychology: Health and Well-Being, 3(1), 1–43. https://doi.org/10.1111/j.1758-0854.2010.01045.x

Dohrmann, D. (2018). Health-Related Perception and Responsibility Behavior Among Individuals With and Without Chronic Illness [University of Nebraska Medical Center]. https://digitalcommons.unmc.edu/cgi/viewcontent.cgi?article=1313&context=etd

Ellis, L. A., Pomare, C., Gillespie, J. A., Root, J., Ansell, J., Holt, J., Wells, L., Tran, Y., Braithwaite, J., & Zurynski, Y. (2021). Changes in public perceptions and experiences of the Australian health care system: A decade of change. Health Expectations, 24(1), 95–110. https://doi.org/10.1111/hex.13154

Fayers, P. M., & Sprangers, M. A. (2002). Understanding self-rated health. The Lancet, 359(9302), 187–188. https://doi.org/10.1016/S0140-6736(02)07466-4

Forte, R., Boreham, C., De Vito, G., & Pesce, C. (2015). Health and Quality of Life Perception in Older Adults: The Joint Role of Cognitive Efficiency and Functional Mobility. International Journal of Environmental Research and Public Health, 12(9), 11328–11344. https://doi.org/10.3390/ijerph120911328

Galenkamp, H., Deeg, D. J. H., Huisman, M., Hervonen, A., Braam, A. W., & Jylha, M. (2013). Is Self-Rated Health Still Sensitive for Changes in Disease and Functioning Among Nonagenarians? The Journals of Gerontology Series B: Psychological Sciences and Social Sciences, 68(5), 848–858. https://doi.org/10.1093/geronb/gbt066

Garrity, T. F., Somes, G. W., & Marx, M. B. (1978). Factors influencing self-assessment of health. Social Science & Medicine. Part A: Medical Psychology & Medical Sociology, 12, 77–81. https://doi.org/10.1016/0271-7123(78)90032-9

Goldstein, M. S., Siegel, J. M., & Boyer, R. (1984). Predicting changes in perceived health status. American Journal of Public Health, 74(6), 611–614. https://doi.org/10.2105/AJPH.74.6.611

Grilli, L., & Rampichini, C. (2014). Ordered Logit Model. In A. C. Michalos (Ed.), Encyclopedia of Quality of Life and Well-Being Research (pp. 4510–4513). Springer Netherlands. https://doi.org/10.1007/978-94-007-0753-5_2023

Ha, J., & Kim, J. (2019). Factors influencing perceived health status among elderly workers: Occupational stress, frailty, sleep quality, and motives for food choices. Clinical Interventions in Aging, Volume 14, 1493–1501. https://doi.org/10.2147/CIA.S210205

Harrington, J., Perry, I. J., Lutomski, J., Fitzgerald, A. P., Shiely, F., McGee, H., Barry, M. M., Van Lente, E., Morgan, K., & Shelley, E. (2010). Living longer and feeling better: Healthy lifestyle, self-rated health, obesity and depression in Ireland. The European Journal of Public Health, 20(1), 91–95. https://doi.org/10.1093/eurpub/ckp102

Husain, Z., & Ghosh, D. (2017). Analysis of Perceived Health Status Among Elderly in India: Gender and Positional Objectivity. In T. Samanta (Ed.), Cross-Cultural and Cross-Disciplinary Perspectives in Social Gerontology (pp. 177–202). Springer Singapore. https://doi.org/10.1007/978-981-10-1654-7_10

Idler, E. L., & Benyamini, Y. (1997). Self-Rated Health and Mortality: A Review of Twenty-Seven Community Studies. Journal of Health and Social Behavior, 38(1), 21. https://doi.org/10.2307/2955359

Jylhä, M. (2009). What is self-rated health and why does it predict mortality? Towards a unified conceptual model. Social Science & Medicine, 69(3), 307–316. https://doi.org/10.1016/j.socscimed.2009.05.013

Kananen, L., Enroth, L., Raitanen, J., Jylhävä, J., Bürkle, A., Moreno-Villanueva, M., Bernhardt, J., Toussaint, O., Grubeck-Loebenstein, B., Malavolta, M., Basso, A., Piacenza, F., Collino, S., Gonos, E. S., Sikora, E., Gradinaru, D., Jansen, E. H. J. M., Dollé, M. E. T., Salmon, M., … Jylhä, M. (2021). Self-rated health in individuals with and without disease is associated with multiple biomarkers representing multiple biological domains. Scientific Reports, 11(1), 6139. https://doi.org/10.1038/s41598-021-85668-7

Krewski, D., Lemyre, L., Turner, M. C., Lee, J. E. C., Dallaire, C., Bouchard, L., Brand, K., & Mercier, P. (2006). Public Perception of Population Health Risks in Canada: Health Hazards and Sources of Information. Human and Ecological Risk Assessment: An International Journal, 12(4), 626–644. https://doi.org/10.1080/10807030600561832

Kumar, S., & Pradhan, M. R. (2019). Self-rated health status and its correlates among the elderly in India. Journal of Public Health, 27(3), 291–299. https://doi.org/10.1007/s10389-018-0960-2

Lee, H.-L., Huang, H.-C., Lee, M.-D., Chen, J. H., & Lin, K.-C. (2012). Factors affecting trajectory patterns of self-rated health (SRH) in an older population—A community-based longitudinal study. Archives of Gerontology and Geriatrics, 54(3), e334–e341. https://doi.org/10.1016/j.archger.2011.10.009

Leinonen, R., Heikkinen, E., & Jylhä, M. (2001). Predictors of decline in self-assessments of health among older people—A 5-year longitudinal study. Social Science & Medicine, 52(9), 1329–1341. https://doi.org/10.1016/S0277-9536(00)00249-5

Lemyre, L., Lee, J. E. C., Mercier, P., Bouchard, L., & Krewski, D. (2006). The structure of Canadians’ health risk perceptions: Environmental, therapeutic and social health risks. Health, Risk & Society, 8(2), 185–195. https://doi.org/10.1080/13698570600677399

Linder, J., McLaren, L., Siou, G. L., Csizmadi, I., & Robson, P. J. (2010). The Epidemiology of Weight Perception: Perceived Versus Self-reported Actual Weight Status among Albertan Adults. Canadian Journal of Public Health, 101(1), 56–60. https://doi.org/10.1007/BF03405563

Machón, M., Vergara, I., Dorronsoro, M., Vrotsou, K., & Larrañaga, I. (2016). Self-perceived health in functionally independent older people: Associated factors. BMC Geriatrics, 16(1), 66. https://doi.org/10.1186/s12877-016-0239-9

May, M. (2006). Cardiovascular disease risk assessment in older women: Can we improve on Framingham? British Women’s Heart and Health prospective cohort study. Heart, 92(10), 1396–1401. https://doi.org/10.1136/hrt.2005.085381

Nair, S., Sawant, N., Thippeswamy, H., & Desai, G. (2021). Gender Issues in the Care of Elderly: A Narrative Review. Indian Journal of Psychological Medicine, 43(5_suppl), S48–S52. https://doi.org/10.1177/02537176211021530

Ranjan, A., & Muraleedharan, V. R. (2020). Equity and elderly health in India: Reflections from 75th round National Sample Survey, 2017–18, amidst the COVID-19 pandemic. Globalization and Health, 16(1), 93. https://doi.org/10.1186/s12992-020-00619-7

Rathbun, K. P., Loerzel, V., & Edwards, J. (2020). Personal Perception of Health in Urban Women of Low Socioeconomic Status: A Qualitative Study. Journal of Primary Care & Community Health, 11, 215013272092595. https://doi.org/10.1177/2150132720925951

Singh, L., Arokiasamy, P., Singh, P. K., & Rai, R. K. (2013). Determinants of Gender Differences in Self-Rated Health Among Older Population: Evidence From India. SAGE Open, 3(2), 215824401348791. https://doi.org/10.1177/2158244013487914

Smith, J. P., Majmundar, M. K., & National Research Council (U.S.) (Eds.). (2012). Aging in Asia: Findings from new and emerging data initiatives. National Academies Press.

UCLA: Statistical Consulting. (2021). ORDERED LOGISTIC REGRESSION. Institute for DIgital Research & Education. https://stats.idre.ucla.edu/stata/output/ordered-logistic-regression/

Wagner, D. C., & Short, J. L. (2014). Longitudinal Predictors of Self-Rated Health and Mortality in Older Adults. Preventing Chronic Disease, 11, 130241. https://doi.org/10.5888/pcd11.130241

Warmoth, K., Tarrant, M., Abraham, C., & Lang, I. A. (2016). Older adults’ perceptions of ageing and their health and functioning: A systematic review of observational studies. Psychology, Health & Medicine, 21(5), 531–550. https://doi.org/10.1080/13548506.2015.1096946

Watcharanat, P., Tanpichai, P., & Sajjasophon, R. (2019). The Relationship Between The Perception of Elderly’s Health Status and Health Behaviors in Nakhon Nayok Province, Thailand. The Open Public Health Journal, 12(1), 420–423. https://doi.org/10.2174/1874944501912010420

Welin, C., Wilhelmsen, L., Welin, L., Johansson, S., & Rosengren, A. (2011). Perceived Health in 50-Year-Old Women and Men and the Correlation With Risk Factors, Diseases, and Symptoms. Gender Medicine, 8(2), 139–149. https://doi.org/10.1016/j.genm.2011.03.005

WHO. (2020). Decade of Healthy Ageing: Plan of Action. https://www.who.int/initiatives/decade-of-healthy-ageing

WHO. (2021). Ageing and health: Fact sheets. https://www.who.int/news-room/fact-sheets/detail/ageing-and-health

Xu, D., Arling, G., & Wang, K. (2019). A cross-sectional study of self-rated health among older adults: A comparison of China and the United States. BMJ Open, 9(7), e027895. https://doi.org/10.1136/bmjopen-2018-027895

Zimmer, Z., Natividad, J., Lin, H.-S., & Chayovan, N. (2000). A Cross-National Examination of the Determinants of Self-Assessed Health. Journal of Health and Social Behavior, 41(4), 465. https://doi.org/10.2307/2676298

